# Vitreous fluid-isolated DNA for the genetic analysis of primary uveal melanoma: a proof-of-concept study

**DOI:** 10.1101/2024.02.09.24302604

**Authors:** R.J. Nell, M. Versluis, N.V. Menger, M.C. Gelmi, T.H.K. Vu, R.M. Verdijk, G.P.M. Luyten, M.J. Jager, P.A. van der Velden

## Abstract

**Background:** Uveal melanoma is an aggressive ocular malignancy. Early molecular characterisation of primary tumours is crucial to identify those at risk of metastatic dissemination. Although tumour biopsies are being taken, liquid biopsies of ocular fluids may form a less invasive but relatively unexplored alternative. In this study, we aim to evaluate the DNA content of vitreous fluid from eyes with a uveal melanoma to obtain molecular information from the tumour.

**Methods:** DNA was isolated from 65 vitreous fluid samples from enucleated eyes with a uveal melanoma and studied using digital PCR. Primary and additional driver mutations (in *GNAQ*, *GNA11*, *PLCB4*, *CYSLTR2*, *BAP1*, *SF3B1* and *EIF1AX*) were investigated using accustomed targeted and drop-off assays. The copy numbers of chromosome 3p and 8q were measured using multiplex and single-nucleotide polymorphism-based assays. Our findings were compared to the molecular profile of matched primary tumours and to the clinicopathological tumour characteristics.

**Results:** Almost all (63/65) vitreous fluids had measurable levels of DNA, but melanoma-cell derived DNA (containing the primary driver mutation) was detected in 39/65 samples (median proportion 18%, range 0.2%-94%) and was associated with a larger tumour prominence, but not with any of the molecular tumour subtypes. Among the vitreous fluids with melanoma-cell derived DNA, not all samples harboured (analysable) other mutations or had sufficient statistical power to measure copy numbers. Still, additional mutations in *BAP1*, *SF3B1* and *EIF1AX* were detected in 13/15 samples and chromosome 3p and 8q copy numbers matched the primary tumour in 19/21 and 18/20 samples, respectively. Collectively, a clinically-relevant molecular classification of the primary tumour could be inferred from 27/65 vitreous fluids.

**Discussion:** This proof-of-concept study shows that substantial amounts of DNA could be detected in vitreous fluids from uveal melanoma patients, including melanoma-cell derived DNA in 60% of the samples. Prognostically-relevant genetic alterations of the primary tumour could be identified in 42% of the patients. A follow-up study is needed to evaluate our approach in a prospective clinical context.

## Introduction

Uveal melanoma is a rare, aggressive intraocular tumour originating from malignantly-transformed melanocytes in the iris, ciliary body or choroid (**Figure 1**). The majority of uveal melanomas are located in the relatively inconspicuous and inaccessible posterior segment of the eye [1]. Consequently, primary tumours can be large upon first detection and (micro)metastases may occur before first treatment [2, 3]. For that reason, early and adequate identification and characterisation of primary uveal melanomas is crucial to prevent tumour progression and ameliorate patient outcome.

**Figure 1.**
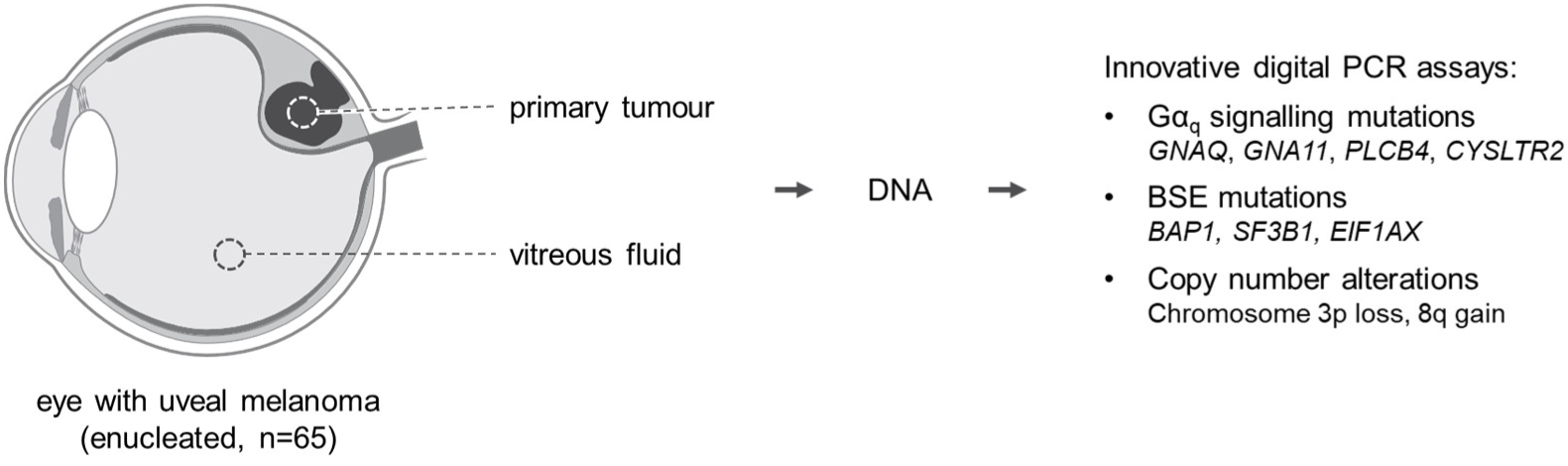
Workflow of current study: DNA from primary tumours [20] and vitreous fluid samples in 65 enucleated eyes is analysed by digital PCR.

Over the past decades, prognostically-relevant molecular tumour subtypes have been identified based on the genetic alterations present in the primary uveal melanoma. Tumours with *EIF1AX* mutations are generally copy number stable and have the best prognosis with the lowest risk of early metastatic dissemination [4, 5]. *SF3B1*-mutant tumours often present with additional copies of chromosome 8q and are associated with an intermediate prognosis with relatively late metastases [4, 6–8]. Uveal melanomas with a loss of chromosome 3p and a *BAP1* mutation, typically also showing chromosome 8q copy number increases, have the poorest prognosis [9, 10].

Primary uveal melanoma is usually treated by local, eye-preserving procedures (brachytherapy, proton beam therapy) or a complete surgical removal of the eye (enucleation) [1]. In case of the latter, tumour tissue is readily available for molecular analysis, and such analysis is usually offered to all enucleated patients. For non-enucleated patients, however, an additional tumour biopsy is required to obtain a molecular profile of the melanoma. While intra-ocular fine-needle aspirate tumour biopsies are being taken routinely in a number of clinics, the procedure is relatively invasive and remains accompanied by various risks [11, 12]. A less invasive (outpatient) biopsy procedure would allow more patients to have a personalised molecular diagnosis and prognosis. Moreover, those with an increased risk of metastatic dissemination may be offered a more intense follow-up screening program, and could possibly benefit from earlier detection of metastasis. Additionally, knowledge about the molecular profile of the primary tumour has been positively associated with various aspects of psychosocial well-being [13] and might be of (future) clinical relevance with regard to novel (neo-)adjuvant therapies [5, 14, 15].

Previously, we introduced digital PCR assays to analyse the key genetic alterations in uveal melanoma to quantify the intratumour heterogeneity in bulk tumour specimens [16–20]. Digital PCR can also be used for the sensitive detection of genetic alterations in scarce DNA samples, such as liquid biopsies (i.e. samples of body fluids such as blood or serum, urine or aspirates) [21]. In this study (**Figure 1**), we isolate DNA from 65 vitreous fluid samples obtained from enucleated eyes with a uveal melanoma. These samples are analysed for mutations in *GNAQ*, *GNA11*, *PLCB4*, *CYSLTR2*, *EIF1AX*, *SF3B1* and *BAP1,* and copy number alterations (CNAs) affecting chromosomes 3p and 8q. Next, our findings are compared to the molecular profile of matched primary tumours and to the clinicopathological tumour characteristics. Collectively, this study forms the proof-of-concept for a comprehensive characterisation of the DNA content of the vitreous as a potential liquid biopsy for uveal melanoma patients to identify those at high risk of developing metastases.

## Materials and methods

### Vitreous fluid sample collection and DNA isolation

Vitreous fluid samples were collected from 65 enucleated eyes with a uveal melanoma as part of the biobank of the Department of Ophthalmology, Leiden University Medical Center (LUMC). Samples had been isolated by syringe suction directly after opening the enucleated eye, and stored at -80 °C. This study was approved by the LUMC Biobank Committee and Medisch Ethische Toetsingscommissie under numbers B14.003/SH/sh and B20.026/KB/kb.

Total DNA was isolated from 100-200 uL vitreous fluid, using the Quick-cfDNA Serum & Plasma Kit (Zymo Research, Irvine, USA) and eluted in 35 uL DNA elution buffer (Zymo), following the manufacturer’s instructions. From all samples, the matched primary uveal melanoma was analysed in our recent study using digital PCR and targeted DNA and RNA sequencing [20]. Clinical and molecular information of all samples is summarised in **Supplementary Table 1**.

### Digital PCR experiments

All digital PCR experiments were carried out using the QX200 Droplet Digital PCR System (Bio-Rad Laboratories, Hercules, USA) using the assays and following the protocols described earlier [16–20, 22]. Typically, 3.5 uL isolated vfDNA was analysed in a single 22 uL experiment, using 11 uL ddPCR™ Supermix for Probes (No dUTP, Bio-Rad) and primers and FAM- or HEX-labelled probes in final concentrations of 900 and 250 nM for duplex experiments, or optimised concentrations for multiplex experiments. Assay details are described in our recent study [20].

All PCR mixtures were partitioned into 20,000 droplets using the AutoDG System (Bio-Rad). Next, a PCR protocol was performed in a T100 Thermal Cycler (Bio-Rad): 10 minutes at 95 °C; 30 seconds at 94 °C and 1 minute at 55 °C, 58 °C or 60 °C for 40 cycles; 10 minutes at 98 °C; cooling at 12 °C for up to 48 hours, until droplet reading. Ramp rate was set to 2 °C/second for all steps.

Reading of the droplets was performed using the QX200 Droplet Reader (Bio-Rad). The raw results of the digital PCR experiments were acquired using *QuantaSoft* (version 1.7.4, Bio-Rad) and imported and analysed in the online digital PCR management and analysis application *Roodcom WebAnalysis* (version 2.0, available via https://webanalysis.roodcom.nl).

First of all, the Gα_q_ signalling mutations (in *GNAQ*, *GNA11*, *CYSLTR2* and *PLCB4*) were analysed in duplex experiments and the measured concentrations (listed between square brackets) of the mutant and wild-type alleles were used to calculate the total vfDNA concentrations and melanoma-cell derived vfDNA fractions:

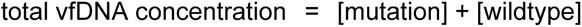

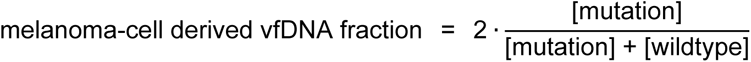

Additional BSE mutations in *BAP1*, *SF3B1* and *EIF1AX* were interpreted in a qualitative manner only.

CNAs were measured via two different approaches [19]. Following the classic approach, copy number values were calculated based on the measured concentrations of a DNA target on chromosome 3p (*PPARG*) and 8q (*PTK2*), and a reference gene on chromosome 5p (*TERT*) or 14q (*TTC5*) assumed to be stable:

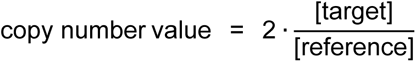

Following the SNP-based approach, copy number values were measured using the concentrations of haploid genomic loci var_1_ and var_2_, representing the two variants of a heterozygous SNP:

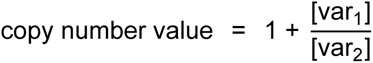

In this formula, the SNP variant in the denominator (var_2_) is the one that was assumed to be unaltered and copy number stable (i.e. haploid). Analysed SNPs in this study included rs6617, rs6976, rs9586, rs1989839, rs1062633 and rs2236947 on chromosome 3p, and rs7018178 and rs7843014 on chromosome 8q.

### In silico evaluation of digital PCR sensitivity and power prediction

To evaluate the statistical power of detecting CNAs in the vfDNA+/UM+ samples, we performed *in silico* digital PCR simulations using our R library *digitalPCRsimulations* (available via https://github.com/rjnell/digitalPCRsimulations), as described recently [19]. In short, for each individual sample, we determined the sensitivity (% of simulated experiments) of detecting a chromosomal loss (copy number = 1) or gain (copy number = 3), assuming that this alteration is clonally present in the entire vfDNA melanoma-cell population. An alteration was considered ‘detected’ when the obtained copy number was lower or higher than 2 and the constructed 99%-confidence interval did not contain 2. The input vfDNA concentration and melanoma-cell fraction were based on the initial Gα_q_ mutation and wild-type measurements. The total amount of accepted droplets was set to 15,000 and each condition was simulated up to 1,000 times. An experimental setup (classic or SNP-based approach) was considered ‘underpowered’ in samples with a sensitivity <80% when analysing a maximum of 45,000 droplets.

### Statistical analysis and code availability

Statistical evaluations in this study were conducted using Spearman’s rank correlation coefficient, Fisher’s exact test, the Mann-Whitney *U* test and the log-rank test. All analyses were performed using R version 4.0.3 in RStudio version 1.4.1103. All custom scripts are via https://github.com/rjnell/um-vitreous.

## Results

### DNA composition of vitreous fluid samples assessed by mutant and wild-type Gα_q_

Gα_q_ signalling mutations at established hotspots in *GNAQ*, *GNA11*, *CYSLTR2* and *PLCB4* are considered the initiating events driving the development of uveal melanoma [18, 23–28]. As they form the most clonal and most common genetic alteration in the primary tumour, they represent a relatively generic marker of the malignant cells that can be used to quantify the abundance of melanoma-cell derived DNA [20]. Based on the exact Gα_q_ mutation that was identified earlier in the matched primary tumour [20], we investigated all vitreous fluid DNA (vfDNA) samples using targeted digital PCR for the absolute and relative abundance of mutant and wild-type DNA (**Figure 2** and **3A**).

**Figure 2.**
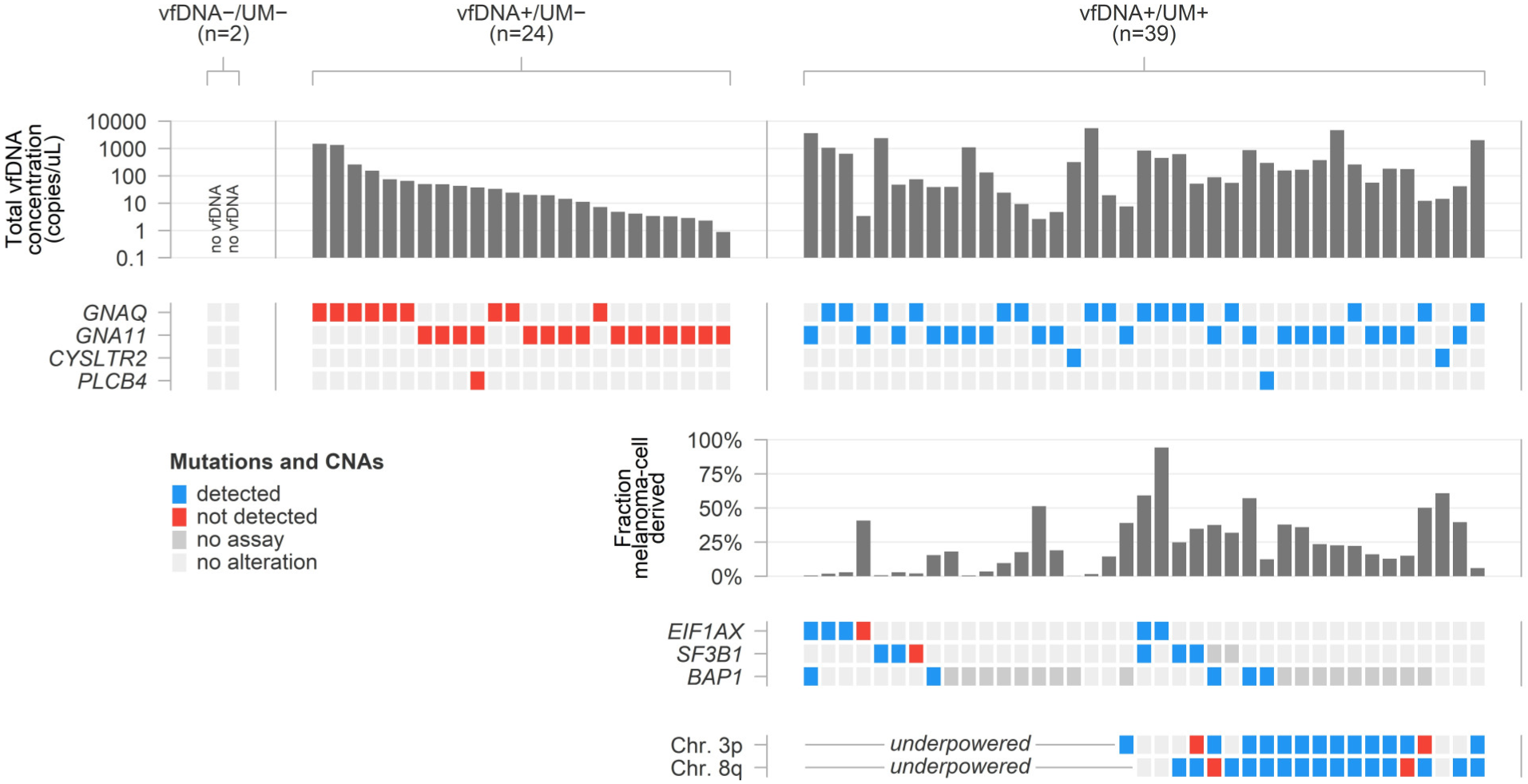
DNA composition and detectability of molecular alterations in 65 vitreous fluid samples. Mutations and CNAs are compared to those identified in the matched primary tumour [20], and their detection in the vfDNA is visualised. ‘No assay’ refers to a selection of mutations that were known to be present in the primary tumour, but could not be analysed in the vfDNA sample. ‘Underpowered’ refers to the condition in which the amount of total and melanoma-cell derived vfDNA was predicted to be insufficient to detect chromosomal CNAs.

**Figure 3.**
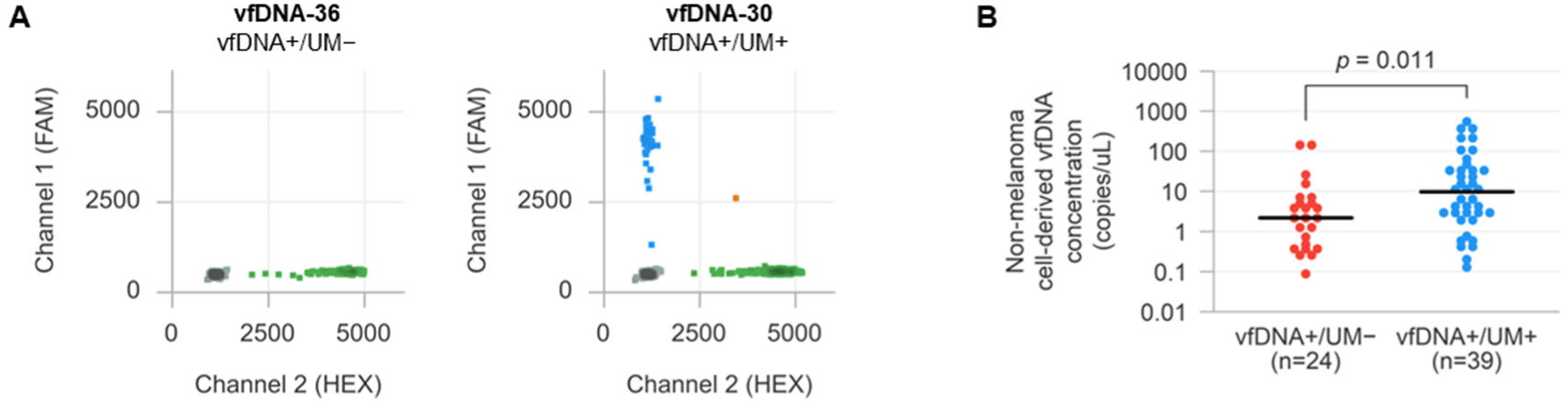
(**A**) Analysis of Gα_q_ signalling mutations using targeted digital PCR, here exemplified for the measurement of *GNA11* p.Q209L mutant and wild-type DNA alleles with two representative 2D plots for a vfDNA+/UM− and vfDNA+/UM+ sample. (**B**) Comparison of non-melanoma cell-derived DNA concentrations in vfDNA+/UM− and vfDNA+/UM+ samples.

We did not find mutant or wild-type alleles in 2/65 (3%) samples, indicating that these samples did not contain measurable amounts of vfDNA (**Figure 2**). Wild-type Gα_q_ alleles only were detected in 24/65 (37%) vitreous fluid samples; these samples were labelled as vfDNA+/UM−, as they had measurable levels of vfDNA but lacked DNA that was derived from mutant melanoma cells (**Figure 2** and **3A**). Still, they contained up to ∼1500 genomic copies per uL vitreous fluid (median 22 genomic copies per uL). In one vfDNA+/UM− patient, the primary uveal melanoma was known to carry two clonally-abundant Gα_q_ signalling mutations (*GNA11* p.R183H and *PLCB4* p.D630N in vfDNA-29), but the vfDNA only contained wild-type *GNA11* and *PLCB4* in comparable concentrations.

Mutant Gα_q_ alleles were detected in 39/65 (60%) samples, which were labelled as vfDNA+/UM+ (**Figure 2** and **3A**). In these samples, the sum of mutant and wild-type allele concentrations was taken as total vfDNA concentration (median 134 genomic copies per uL). The mutant allele fractions were used to estimate the abundance of melanoma-cell derived vfDNA, revealing proportions between 0.2% and 94% (median 18%). vfDNA+/UM+ samples also contained significantly more non-melanoma cell derived vfDNA (i.e. DNA not derived from uveal melanoma cells, Mann-Whitney U *p* = 0.011) than vfDNA+/UM− samples (**Figure 3B**).

### Analysis of additional BSE mutations in *BAP1*, *SF3B1* and *EIF1AX*

Besides the generic Gα_q_ mutations, the majority of all uveal melanomas are known to carry an additional ‘BSE mutation’ in *BAP1*, *SF3B1* or *EIF1AX* [4, 7, 9]. In contrast to the Gα_q_ mutations, however, only a subset of those BSE mutations can be found at a hotspot position. In our cohort of primary uveal melanomas from vfDNA+/UM+ patients [20], *BAP1* alterations occurred throughout the entire gene, most of the *SF3B1* mutations affected position p.R625, and *EIF1AX* mutations were found at various positions within the first two exons of the gene (**Supplementary Table 1**).

To be able to analyse these diverse BSE mutations, we developed a variety of targeted and generic drop-off digital PCR assays [20]. Hereby, a small number of unique *BAP1* alterations, all hotspot mutations in *SF3B1* (p.R625C/H), and all mutations in *EIF1AX* (exon 1 and exon 2) could be analysed (**Figure 4 and Supplementary Table 1**). We now applied these assays in the corresponding cohort of vfDNA+/UM+ samples (n=15). A selection of vfDNA+/UM− samples with matched total vfDNA concentrations were measured as negative controls (**Figure 4**). Based on the known mutational status of the respective primary tumours, *EIF1AX* mutations were successfully identified in 5/6 vitreous fluid samples, *SF3B1* mutations in 5/6 samples and *BAP1* mutations in 5/5 samples (**Figure 2**). Two samples (vfDNA-07 and vfDNA-56) carried two BSE mutations, which were both detected in the vfDNA. Of note, the two vitreous fluid samples with undetected BSE mutations (vfDNA-23 and vfDNA-49) had two of the lowest concentrations of Gα_q_ mutant alleles within the vfDNA+/UM+ cohort (**Figure 2 and Supplementary Table 1**). In total, 13/15 (87%) of the analysed vitreous fluid samples had a measurable BSE mutation matching with the corresponding primary tumour.

**Figure 4.**
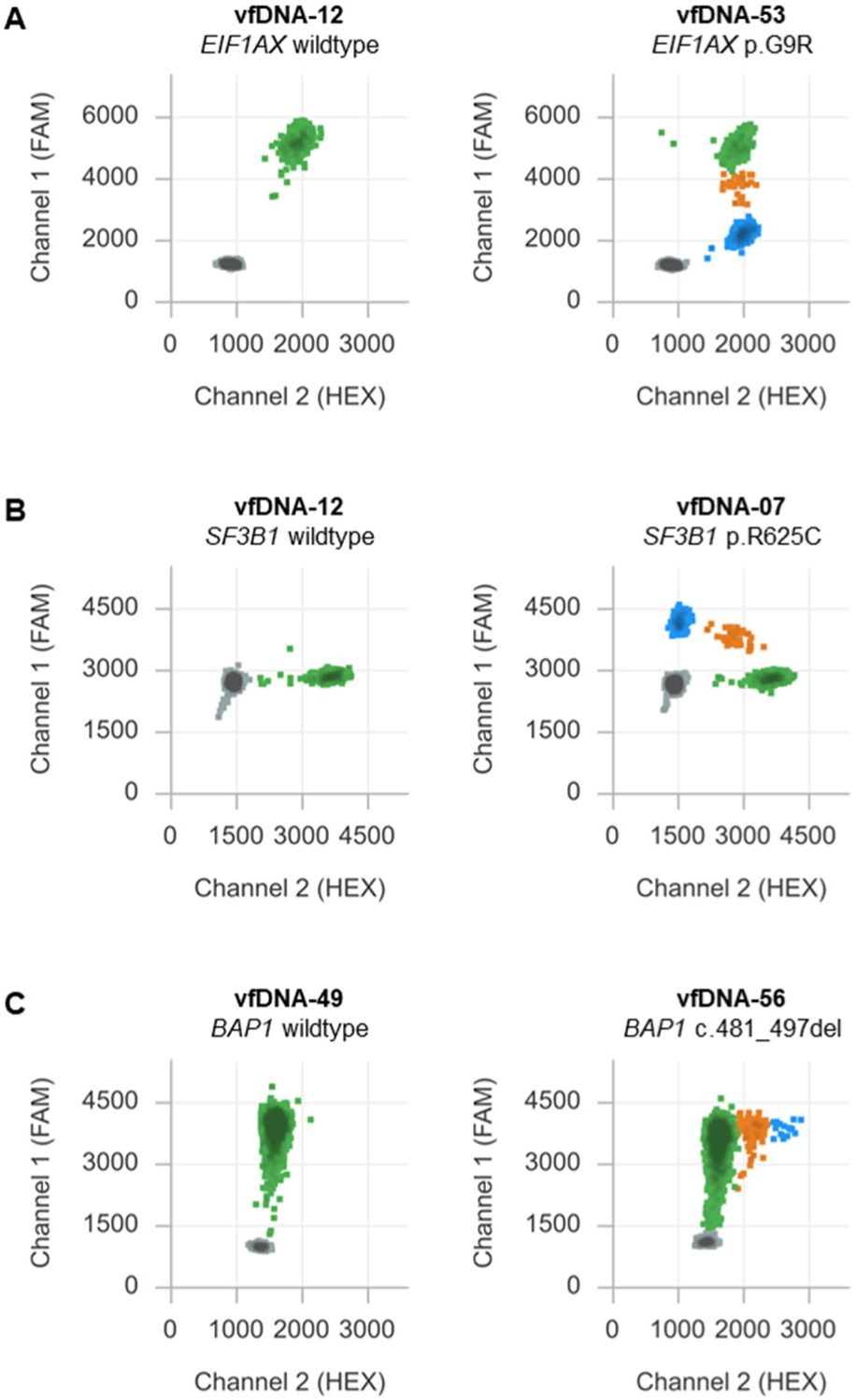
Analysis of BSE mutations in *EIF1AX* (**A**), *SF3B1* (**B**) and *BAP1* (**C**) using drop-off and targeted digital PCR assays. Here exemplified with the representative 2D plots for a wild-type and mutant vfDNA sample with similar total vfDNA concentrations.

### Advanced chromosome 3p and 8q copy number analysis

CNAs are usually studied by comparing the absolute abundance of a target of interest to a stable genomic reference (classic approach, **Figure 5A**) [19]. As an alternative, CNAs may be measured by evaluating the allelic (im)balance between the two variants of a heterozygous SNP on the chromosome of interest (SNP-based approach, **Figure 5A**). While both SNP variants are equally abundant under healthy conditions, this balance is altered when there is a copy number loss or gain of one of the alleles. Conveniently, a stable genomic reference is not required when using this approach and more precise measurements can be obtained. We recently validated this methodology for detecting chromosome 3p and 8q alterations in uveal melanoma [19].

**Figure 5.**
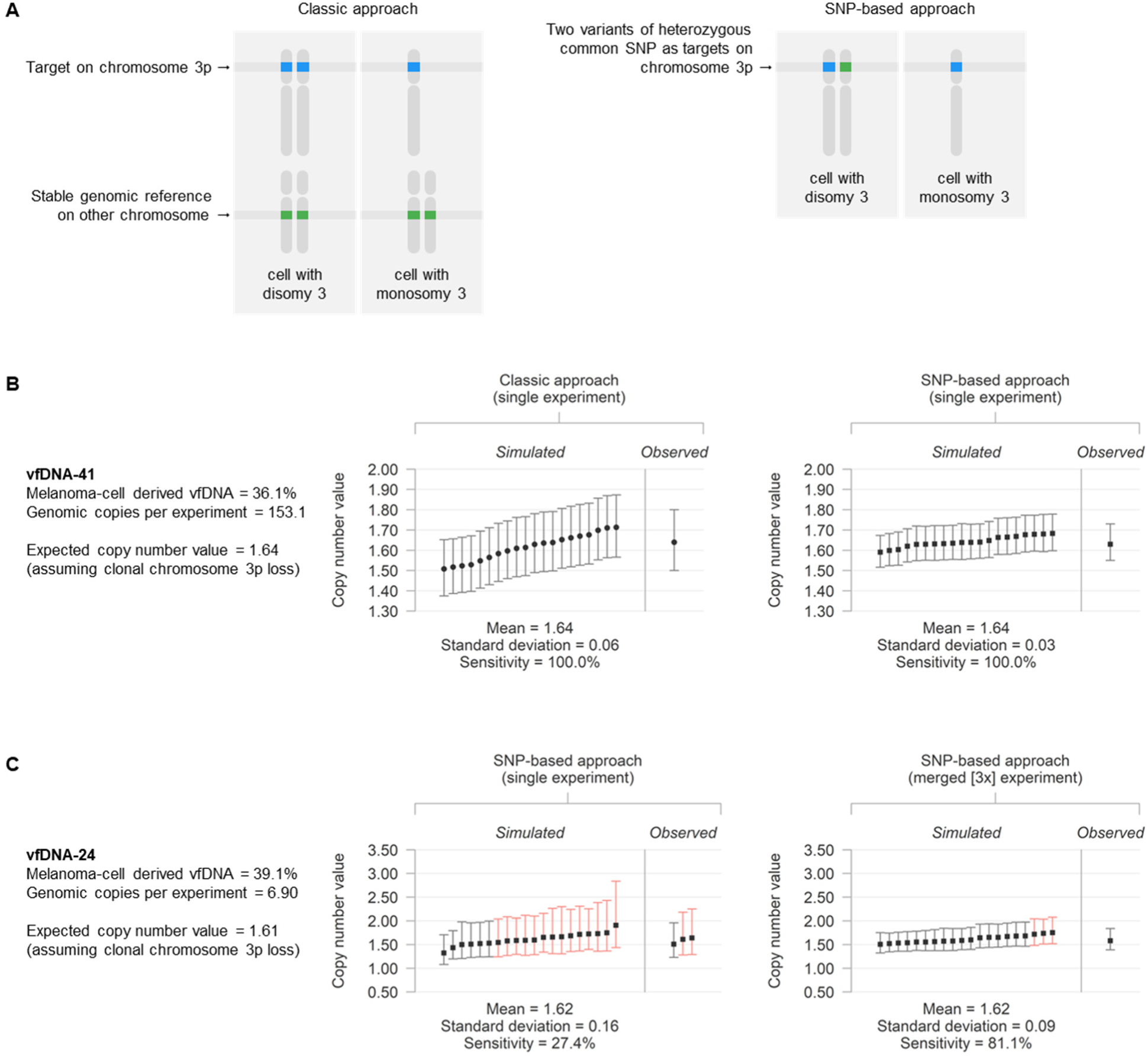
(**A**) Approaches to analyse copy number alterations, here exemplified for measuring chromosome 3p loss. (**B**) Simulated and observed copy number value measurements for chromosome 3p in vfDNA-41, with associated confidence intervals. Simulations were based on the initial Gα_q_ mutant and wild-type measurements and the assumption of a clonal loss in the vfDNA melanoma-cell population, in single experiments using both the classic and SNP-based approach. Significance of the measurements is indicated by the colour of the individual confidence interval (red: not significantly different from 2 [*p* > 0.01], grey: significantly different from 2 [*p* < 0.01]). (**C**) Overview of simulated and observed copy number value measurements for chromosome 3p in vfDNA-24, in both single and (3x) merged experiments using the SNP-based approach.

The precision of digital PCR measurements also depends on the amount of DNA input (limited by the low quantity of our samples) and the number of partitions (i.e. droplets) [19]. A typical digital PCR experiment yields between 10,000 and 20,000 droplets, but this can be increased by repeating the experiment and merging the results into one combined analysis. The limiting factor, however, remains that those experimental replicates also require more sample DNA. To determine the statistical power of both the classic and SNP-based approach in our application of analysing samples with diverse total and melanoma-cell derived vfDNA concentrations, we performed *in silico* simulations based on the initial Gα_q_ mutant and wild-type measurements of all vfDNA+/UM+ samples. For each individual sample, we evaluated the sensitivity of detecting a chromosomal loss (copy number = 1) or gain (copy number = 3) assuming that this alteration is clonally present in the vfDNA melanoma-cell population. As representative examples, the *in silico* analyses and *in vitro* observations for measuring chromosome 3p loss in samples vfDNA-41 and vfDNA-24 are presented in **Figure 5B and C**. A summary of the analyses for all samples is given in **Supplementary Table 1**.

Some vfDNA samples were found to contain sufficient amounts of DNA to detect a chromosomal alteration in a single digital PCR experiment (such as vfDNA-41 in **Figure 5B**). In other samples, the sensitivity of detecting a CNA was lower, but this could be increased by repeating the experiment up to three times and merging the results (such as vfDNA-24 in **Figure 5C**). In general, higher levels of sensitivity were obtained when more droplets could be analysed, when more melanoma-cell derived vfDNA was present and when less healthy cell DNA ‘contaminated’ the sample (**Supplementary Table 1**). Moreover, in line with our earlier observations [19, 20], the SNP-based approach was generally more precise than the classic alternative (**Figure 5C**) and was chosen as preferred methodology whenever available.

Based on the *in silico* results, we limited further copy number analysis to those vitreous fluid samples with an expected sensitivity >80% of calling a loss (of chromosome 3p, in 21/39 = 54% of the vfDNA+/UM+ samples) or gain (of chromosome 8q, in 20/39 = 51% of the vfDNA+/UM+ samples) in the available amount of vfDNA (**Materials and methods and Figure 2**). Within this subgroup, chromosome 3p losses were detected in 13/15 samples with a known loss in the matched primary tumour. Similarly, chromosome 8q copy number increases were detected in 15/17 samples with a known chromosome 8q CNA in the tumour. It is noteworthy that in all four discordant cases, the CNA (that remained undetected in the vfDNA) was heterogeneously present in the matched primary tumour: two uveal melanomas had a subclonal loss of chromosome 3p and two a subclonal gain of chromosome 8q (**Supplementary Table 1**) [20]. We did not detect CNAs in vfDNA+/UM+ samples with no chromosome 3p (n=6) or 8q (n=3) alterations in the matched primary tumour, nor in a selection of underpowered vfDNA+/UM+ samples and vfDNA+/UM− samples (which lack copy number altered melanoma-cell derived DNA). Overall, the presence of chromosome 3p and 8q alterations matched those in the primary tumour in 19/21 (90%) and 18/20 (90%) analysable vitreous fluid samples, respectively.

### Clinicopathological context

Lastly, we investigated whether the vfDNA content correlated to the clinicopathological context of the primary tumour. First of all, we hypothesised that the detectability of melanoma-cell derived vfDNA related to anatomical characteristics of the primary tumour (**Figure 6A and B**). No significant differences were found with regard to the largest basal diameter and frequency tumours broken through Bruch’s membrane, but patients with melanoma-cell derived vfDNA had a larger prominence of their primary tumour (median 8 versus 6 mm, Mann-Whitney U *p* = 0.018) than those without. Within the vfDNA+/UM+ samples, there was a significant positive correlation between the prominence of primary tumour and concentration of melanoma-cell derived vfDNA (Spearman’s rho = 0.52, *p* < 0.001, **Figure 6C**).

**Figure 6.**
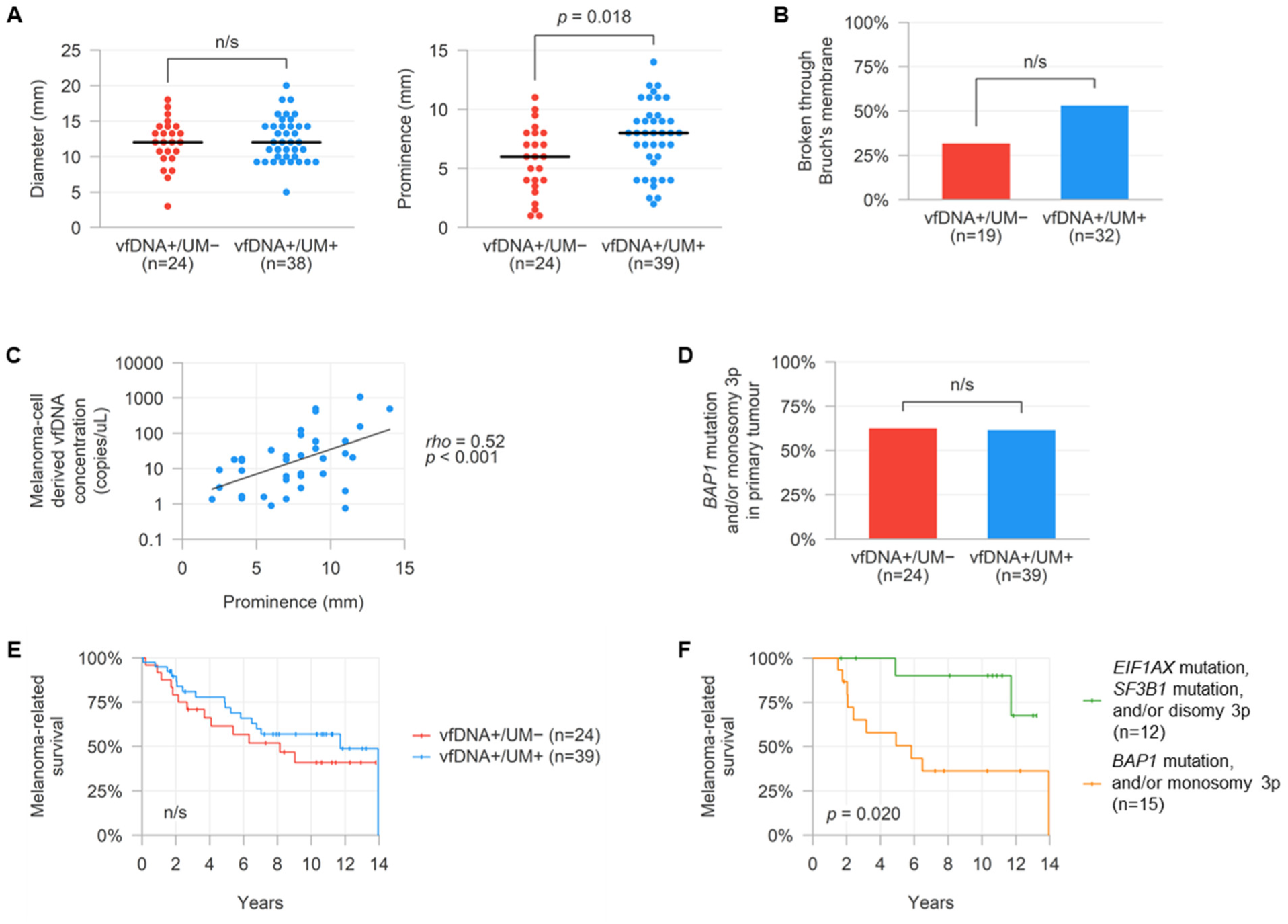
(**A**) Comparison in pathologically-determined largest basal diameter and prominence of matched primary tumours in vfDNA+/UM− or UM+ samples. The black horizontal line denotes the median value for both groups. (**B**) Comparison in frequency of pathologically-determined breaching of Bruch’s membrane in matched primary tumours in vfDNA+/UM+ and UM− samples. (**C**) Correlation between concentrations of mutant vfDNA and pathologically-determined prominence of matched primary tumours in vfDNA+/UM+ samples. (**D**) Comparison in frequency of matched primary tumours with a poor prognostic molecular subtype (*BAP1* mutation and/or monosomy 3p) in vfDNA+/UM− or UM+ samples. (**E**) Melanoma-related survival of patients within the vfDNA+/UM− or UM+ groups. (**F**) Melanoma-related survival of patients based on the inferred molecular subtypes from vfDNA measurements.

Based on the genetic analysis of the primary tumours [20], the distribution of the various molecular uveal melanoma subtypes within the vfDNA+/UM− and vfDNA+/UM+ patients was found to be very similar (**Figure 6D and Supplementary Table 1**). Not unexpectedly, the absence or presence of melanoma-cell derived vfDNA itself did not show an association with melanoma-related survival (**Figure 6E**). Focussing on the subsequent vfDNA measurements, however, the molecular subtype of the primary tumour could be successfully inferred from 27/65 (42%) of the vfDNA samples. Within this subgroup, the prognostic relevance of the molecular classification was evident (log-rank *p* = 0.020, **Figure 6F**): patients with a *BAP1* mutation and/or monosomy 3p had a median melanoma-related survival of 5.8 years, while the estimated median survival was not reached in the group of patients with an *EIF1AX* mutation, *SF3B1* mutation and/or disomy 3p.

## Discussion

In various malignancies, the analysis of liquid biopsies has proven to provide clinically-relevant molecular information of primary or metastatic tumours, while forming a less invasive alternative to direct tumour biopsies [21, 29]. In ocular oncology, the genetic analysis of eye fluids has a prominent role in the differentiation between a malignant intra-ocular lymphoma and a benign uveitis [30], but it remained unexplored in the context of characterising uveal melanoma. For this reason, we investigated the DNA from the vitreous fluid as a potential source of molecular information from the primary tumour (**Figure 1**).

Firstly, we measured the total amount and fraction of melanoma-cell derived vfDNA based on the abundance of mutant and wild-type Gα_q_ alleles (**Figure 2 and 3**). This analysis was successful in 97% of the samples, indicating that nearly all vitreous fluids contained measurable levels of DNA. However, in only 60% of all samples part of this genetic material was derived from melanoma cells. This indicates that a considerable proportion of the DNA present in the vitreous originated from non-malignant cells. It would be interesting to further characterise the cell type of origin of this DNA, for example by applying genetic immune cell measurements or epigenetic profiling [31–34].

Next, we tested the vfDNA+/UM+ samples for additional DNA alterations. Based on the known genetic profile of the matched primary tumour, 13/15 (87%) of the analysed vitreous fluid samples had a measurable BSE mutation (**Figure 2 and 4**). Importantly, however, this supervised mutational screening is not feasible when no matched tumour material is available. In such situations, the vfDNA should be screened for all possible alterations, but this procedure requires multiple experiments and is sample-expensive. A possible solution may be found in the development of multiplex setups in which various assays are combined within one experiment. Our *EIF1AX* drop-off assays furthermore proved that mutation analyses can be made more generic: all different exon 1 and 2 mutations could be measured requiring only two distinct assays. Similar drop-off approaches may be developed for the Gα_q_ signalling mutations and the majority of *SF3B1* mutations. However, it is unlikely that comparable assays can be developed to generically detect all *BAP1* mutations, as these mutations do no occur at hotspot positions and are practically unique in each mutant tumour.

In addition to the BSE mutations, CNAs were studied in the vfDNA+/UM+ samples. The presence of chromosome 3p and 8q alterations corresponded with those in the primary tumour in 19/21 (90%) and 18/20 (90%) of the analysable cases, respectively (**Figure 2 and 5**). Though, we noted that remarkable differences exist in the detectability of melanoma-cell derived mutations versus CNAs: whereas the qualitative identification of any mutant allele inevitably confirms the presence of this mutation, a CNA can only be detected upon a precise quantification of the chromosomal target in comparison to the healthy copy number invariant situation. This leads to sample-specific detection limits that are higher than those for mutations, especially in samples with large amounts of ‘contaminating’ non-melanoma cell derived unaltered DNA. In this study, we first evaluated the detectability of CNAs *in silico*, based on the varying concentrations of (non-)melanoma-cell derived DNA of each individual sample (**Figure 5**). As a result, roughly half of the vfDNA+/UM+ samples were found to have a too low total DNA concentration or fraction melanoma-cell derived DNA, and were technically underpowered to detect chromosomal gains or losses. While this means that no conclusions can be drawn on the presence of CNAs in these samples, this *in silico* analysis helped to avoid generating false-negative results and prevented an unnecessary waste of precious material.

Although most of our vfDNA analyses successfully identified the BSE mutation or CNA present in the primary tumour, a number of cases was discordant (**Figure 2 and Supplementary Table 1**). We noted that the two BSE-lacking vitreous fluid samples presented extreme low purity (< 0.8 Gα_q_ mutant copies/uL), which may challenge the identification of additional mutations with alternative assays. It would be interesting to further compare all digital PCR mutation assays for their exact lower limits of detection, and to repeat experiments to exclude vfDNA subsampling and improve the sensitivity of the mutation detection. In contrast, the four CNA-lacking vitreous fluid samples were predicted to have sufficient melanoma-cell derived DNA concentrations to detect chromosomal gains and losses, assuming that the alteration clonally abundant in the entire melanoma-cell derived fraction vfDNA. Intriguingly, all CNA-discordant cases demonstrated heterogeneity for this chromosomal loss or gain in the primary tumour. This admixed presence of clones with and without a certain alteration may explain why it could not be detected in the vitreous fluid: it may be not present or only present in an undetectable low proportion in the fraction melanoma-cell derived vfDNA.

From a clinical perspective, we found that the detectability of melanoma-cell derived vfDNA was associated with a higher amount of non-tumour cell derived vfDNA and a larger prominence of the primary tumour (**Figure 6A and C**). This suggests that larger (and possibly more invasive) growth of uveal melanomas contributes to the presence of DNA in the vitreous fluid, originating from both melanoma and other cells. However, the identification of melanoma-cell derived vfDNA itself did not mark a genetically distinct or prognostically-relevant subgroup of patients (**Figure 6D and E**). By combining the BSE mutation and CNA results, in 27/65 (42%) of the cases the molecular subtype of the primary tumour could be successfully inferred from the vfDNA measurements. Within this subgroup, the correlation between this molecular classification of tumours and patients’ risk of developing lethal metastases was clearly demonstrated (**Figure 6F**).

Our findings illustrate that in a proportion of patients, relevant molecular information from their primary uveal melanoma may be derived from analysing the DNA present in the vitreous fluid. Following this approach, the risk of metastatic tumour dissemination may be estimated without a direct tumour biopsy, and an increasing number of patients may be offered concordant clinical follow-up. Nevertheless, it is crucial to note that in our current study the vitreous fluid samples were investigated that had been collected ex vivo from enucleated eyes. This procedure is different to it would be in a clinical setting, and uveal melanomas treated by enucleation are typically larger and more invasive than irradiated tumours. Therefore, a follow-up study should evaluate the applicability of our methodology in a prospective clinical context, for example by defining patient groups most likely to benefit from the analysis (e.g. those with relatively thick tumours), and by evaluating vfDNA collected in a similar way as performed in vivo from non-enucleated eyes.

One may also consider to explore possible additions and alternatives to our current methodologies. For example, epigenetic targets (such as DNA methylation) may be evaluated as markers for the various molecular subtypes of uveal melanomas. Candidate loci have already been identified [35, 36], and as DNA methylation can be measured using digital PCR as well [37, 38], the analysis may be easily incorporated in our current workflow. Additionally, the experimental performance might be optimised to reach lower limits of detection, for example by denaturating the DNA samples prior to digital PCR analysis [39]. As further alternatives, vitreous fluid samples may be studied using complementary molecular techniques (such as ultradeep sequencing) or by analysing its RNA, as we recently showed that tumour-derived genetic information may also be inferred from a transcriptional level [40].

Lastly, we believe that our digital PCR assays and approaches have translational potential in the analysis of heterogeneous solid or liquid uveal melanoma biopsies of any other origin. Some of the mutation assays described in this study have already been used in blood-based screening of uveal melanoma patients [41], and the value of similar applications is increasingly recognised [42].

In conclusion, our findings demonstrate the possibility of capturing the molecular profile of a primary uveal melanoma by analysing the DNA present in the vitreous fluid of the eye. As this approach does not require a biopsy to be taken from the tumour itself, a molecular classification of the primary tumour and concordant follow-up might be offered to more patients. In the end, this may ameliorate the clinical outcome of patients with this aggressive malignancy.

## Data Availability

All data produced in the present work are contained in the manuscript.

https://github.com/rjnell/um-vitreous

## Acknowledgements

This study was supported by the European Union’s Horizon 2020 research and innovation program under grant agreement number 667787 (UM Cure 2020, R.J. Nell and N.V. Menger). The authors thank the UM Cure 2020 consortium partners and patient organisations for their helpful discussions regarding this research project.

## Supplementary information

### Supplementary Table 1

Overview of clinical and molecular information of all cases analysed in this study.

## References

1. Jager, M.J., et al., Uveal melanoma. Nat Rev Dis Primers, 2020. 6(1): p. 24.

2. Eskelin, S., et al., Tumor doubling times in metastatic malignant melanoma of the uvea: tumor progression before and after treatment. Ophthalmology, 2000. 107(8): p. 1443–9.

3. Uner, O.E., et al., Estimation of the timing of BAP1 mutation in uveal melanoma progression. Sci Rep, 2021. 11(1): p. 8923.

4. Martin, M., et al., Exome sequencing identifies recurrent somatic mutations in EIF1AX and SF3B1 in uveal melanoma with disomy 3. Nat Genet, 2013. 45(8): p. 933–6.

5. Rodrigues, M., et al., Evolutionary Routes in Metastatic Uveal Melanomas Depend on MBD4 Alterations. Clin Cancer Res, 2019. 25(18): p. 5513–5524.

6. Furney, S.J., et al., SF3B1 mutations are associated with alternative splicing in uveal melanoma. Cancer Discov, 2013. 3(10): p. 1122–1129.

7. Harbour, J.W., et al., Recurrent mutations at codon 625 of the splicing factor SF3B1 in uveal melanoma. Nat Genet, 2013. 45(2): p. 133–5.

8. Yavuzyigitoglu, S., et al., Uveal Melanomas with SF3B1 Mutations: A Distinct Subclass Associated with Late-Onset Metastases. Ophthalmology, 2016. 123(5): p. 1118–28.

9. Harbour, J.W., et al., Frequent mutation of BAP1 in metastasizing uveal melanomas. Science, 2010. 330(6009): p. 1410-3.

10. Robertson, A.G., et al., Integrative Analysis Identifies Four Molecular and Clinical Subsets in Uveal Melanoma. Cancer Cell, 2017. 32(2): p. 204–220 e15.

11. Singh, A.D., et al., Fine-needle aspiration biopsy of uveal melanoma: outcomes and complications. Br J Ophthalmol, 2016. 100(4): p. 456–62.

12. Sellam, A., et al., Fine Needle Aspiration Biopsy in Uveal Melanoma: Technique, Complications, and Outcomes. Am J Ophthalmol, 2016. 162: p. 28–34 e1.

13. Lieb, M., et al., Psychosocial impact of prognostic genetic testing in uveal melanoma patients: a controlled prospective clinical observational study. BMC Psychol, 2020. 8(1): p. 8.

14. Damato, B.E., et al., Tebentafusp: T Cell Redirection for the Treatment of Metastatic Uveal Melanoma. Cancers (Basel), 2019. 11(7).

15. Bigot, J., et al., Splicing Patterns in SF3B1-Mutated Uveal Melanoma Generate Shared Immunogenic Tumor-Specific Neoepitopes. Cancer Discov, 2021. 11(8): p. 1938–1951.

16. Versluis, M., et al., Digital PCR validates 8q dosage as prognostic tool in uveal melanoma. PLoS One, 2015. 10(3): p. e0116371.

17. de Lange, M.J., et al., Heterogeneity revealed by integrated genomic analysis uncovers a molecular switch in malignant uveal melanoma. Oncotarget, 2015. 6(35): p. 37824–35.

18. Nell, R.J., et al., Involvement of mutant and wild-type CYSLTR2 in the development and progression of uveal nevi and melanoma. BMC Cancer, 2021. 21(1): p. 164.

19. Nell, R.J., et al., Allele-specific digital PCR enhances precision and sensitivity in the detection and quantification of copy number alterations in heterogeneous DNA samples: an in silico and in vitro validation study. medRxiv, 2023.

20. Nell, R.J., et al., Digital PCR-based deep quantitative profiling delineates heterogeneity and evolution of uveal melanoma. medRxiv, 2024.

21. Olmedillas-Lopez, S., M. Garcia-Arranz, and D. Garcia-Olmo, Current and Emerging Applications of Droplet Digital PCR in Oncology. Mol Diagn Ther, 2017. 21(5): p. 493–510.

22. Zoutman, W.H., R.J. Nell, and P.A. van der Velden, Usage of Droplet Digital PCR (ddPCR) Assays for T Cell Quantification in Cancer. Methods Mol Biol, 2019. 1884: p. 1–14.

23. Van Raamsdonk, C.D., et al., Frequent somatic mutations of GNAQ in uveal melanoma and blue naevi. Nature, 2009. 457(7229): p. 599–602.

24. Van Raamsdonk, C.D., et al., Mutations in GNA11 in uveal melanoma. N Engl J Med, 2010. 363(23): p. 2191–9.

25. Johansson, P., et al., Deep sequencing of uveal melanoma identifies a recurrent mutation in PLCB4. Oncotarget, 2016. 7(4): p. 4624–31.

26. Moore, A.R., et al., Recurrent activating mutations of G-protein-coupled receptor CYSLTR2 in uveal melanoma. Nat Genet, 2016. 48(6): p. 675–80.

27. Vader, M.J.C., et al., GNAQ and GNA11 mutations and downstream YAP activation in choroidal nevi. Br J Cancer, 2017. 117(6): p. 884–887.

28. Durante, M.A., et al., Genomic evolution of uveal melanoma arising in ocular melanocytosis. Cold Spring Harb Mol Case Stud, 2019. 5(4).

29. Lone, S.N., et al., Liquid biopsy: a step closer to transform diagnosis, prognosis and future of cancer treatments. Mol Cancer, 2022. 21(1): p. 79.

30. de Hoog, J., et al., Combined cellular and soluble mediator analysis for improved diagnosis of vitreoretinal lymphoma. Acta Ophthalmol, 2019. 97(6): p. 626–632.

31. Zoutman, W.H., et al., Accurate Quantification of T Cells by Measuring Loss of Germline T-Cell Receptor Loci with Generic Single Duplex Droplet Digital PCR Assays. J Mol Diagn, 2017. 19(2): p. 236–243.

32. de Lange, M.J., et al., Digital PCR-Based T-cell Quantification-Assisted Deconvolution of the Microenvironment Reveals that Activated Macrophages Drive Tumor Inflammation in Uveal Melanoma. Mol Cancer Res, 2018. 16(12): p. 1902–1911.

33. Zoutman, W.H., et al., A novel digital PCR-based method to quantify (switched) B cells reveals the extent of allelic involvement in different recombination processes in the IGH locus. Mol Immunol, 2022. 145: p. 109–123.

34. van de Leemkolk, F.E.M., et al., Quantification of Unmethylated Insulin DNA Using Methylation Sensitive Restriction Enzyme Digital Polymerase Chain Reaction. Transpl Int, 2022. 35: p. 10167.

35. Field, M.G., et al., BAP1 Loss Is Associated with DNA Methylomic Repatterning in Highly Aggressive Class 2 Uveal Melanomas. Clin Cancer Res, 2019. 25(18): p. 5663–5673.

36. Bakhoum, M.F., et al., BAP1 methylation: a prognostic marker of uveal melanoma metastasis. NPJ Precis Oncol, 2021. 5(1): p. 89.

37. Nell, R.J., et al., Quantification of DNA methylation independent of sodium bisulfite conversion using methylation-sensitive restriction enzymes and digital PCR. Hum Mutat, 2020. 41(12): p. 2205–2216.

38. Salgado, C., et al., Interplay between TERT promoter mutations and methylation culminates in chromatin accessibility and TERT expression. PLoS One, 2020. 15(4): p. e0231418.

39. Fitarelli-Kiehl, M., et al., Denaturation-Enhanced Droplet Digital PCR for Liquid Biopsies. Clin Chem, 2018. 64(12): p. 1762–1771.

40. Nell, R.J., et al., Identification of clinically-relevant genetic alterations in uveal melanoma using RNA sequencing. medRxiv, 2023.

41. Beasley, A., et al., Clinical Application of Circulating Tumor Cells and Circulating Tumor DNA in Uveal Melanoma. JCO Precis Oncol, 2018. 2.

42. Carvajal, R.D., et al., Clinical and molecular response to tebentafusp in previously treated patients with metastatic uveal melanoma: a phase 2 trial. Nat Med, 2022. 28(11): p. 2364–2373.

